# SARS-CoV-2 infection and reinfection in a seroepidemiological workplace cohort in the United States

**DOI:** 10.1101/2021.05.04.21256609

**Authors:** Emilie Finch, Rachel Lowe, Stephanie Fischinger, Michael de St Aubin, Sameed M. Siddiqui, Diana Dayal, Michael A. Loesche, Justin Rhee, Samuel Berger, Yiyuan Hu, Matthew J. Gluck, Benjamin Mormann, Mohammad A. Hasdianda, Elon R. Musk, Galit Alter, Anil S. Menon, Eric J. Nilles, Adam J. Kucharski, on behalf of the CMMID COVID-19 working group and the SpaceX COVID-19 Cohort Collaborative

**Author notes:** Equal contribution (Anil S. Menon, Eric J. Nilles, Adam J. Kucharski).

## Abstract

Identifying the extent of SARS-CoV-2 reinfection is crucial for understanding possible long-term epidemic dynamics. We analysed longitudinal PCR and serological testing data from a prospective cohort of 4411 US employees in four states between April 2020 and February 2021. We conducted a multivariable logistic regression investigating the association between baseline serological status and subsequent PCR test result in order to calculate an odds ratio for reinfection. We estimated an adjusted odds ratio of 0.09 (95% CI: 0.005 – 0.48) for reinfection, implying that the presence of SARS-CoV-2 antibodies at baseline is associated with around 91% reduced odds of a subsequent PCR positive test. This suggests that primary infection with SARS-CoV-2 provides protection against reinfection in the majority of individuals, at least over a sixth month time period. We also highlight two major sources of bias and uncertainty to be considered when estimating reinfection risk, confounders and the choice of baseline time point, and show how to account for both in our analysis.

## Introduction

The rapid global spread of COVID-19 throughout 2020 occurred as a result of the introduction of a highly transmissible virus, SARS-CoV-2, into populations with little pre-existing immunity [1]. Identifying the extent and duration of protective immunity afforded by natural infection (and by vaccination) is therefore of crucial importance for understanding possible long-term epidemic dynamics of SARS-CoV-2 [2].

Studies have estimated that over 95% of symptomatic COVID-19 cases develop antibodies against SARS-CoV-2, with most individuals developing antibodies within three weeks of symptom onset [3,4]. Several serological studies have also characterised individual-level immune dynamics, with some finding evidence for antibody waning and others for sustained antibody responses over several months [5–10]. Antibody kinetics are thought to vary between individuals and are possibly associated with severity of illness, where asymptomatic or mildly symptomatic individuals may develop lower levels of antibodies that wane more rapidly [3,7,11]. While neutralising antibodies are thought to be associated with protection from reinfection, there is currently no established correlate between antibody levels and protective immunity, and the impact of putative antibody waning on reinfection risk is still unclear.

Confirmed cases of reinfection with SARS-CoV-2 have been reported since August 2020 [12]. However, existing large studies examining the relative risk of reinfection in antibody positive individuals have typically involved specific cohorts who may not be representative of the wider community, such as closed communities or healthcare worker cohorts [13–16]. To evaluate the risk of SARS-CoV-2 infection and reinfection over time, we analysed PCR and serological testing data from a prospective cohort of SpaceX employees in the USA between April 2020 and February 2021 [17].

## Results

Of 4411 individuals enrolled, 309 individuals tested seropositive during the study period (Figure 1). This resulted in an overall adjusted seroprevalence of 8.2% (95% CI: 7.3-9.1). We defined a possible reinfection as a new positive PCR test more than 30 days after initial seropositive result. This identified 14 possible reinfections with a median time of 66·5 days between initial seropositive test and PCR positive test.

**Figure 1:**
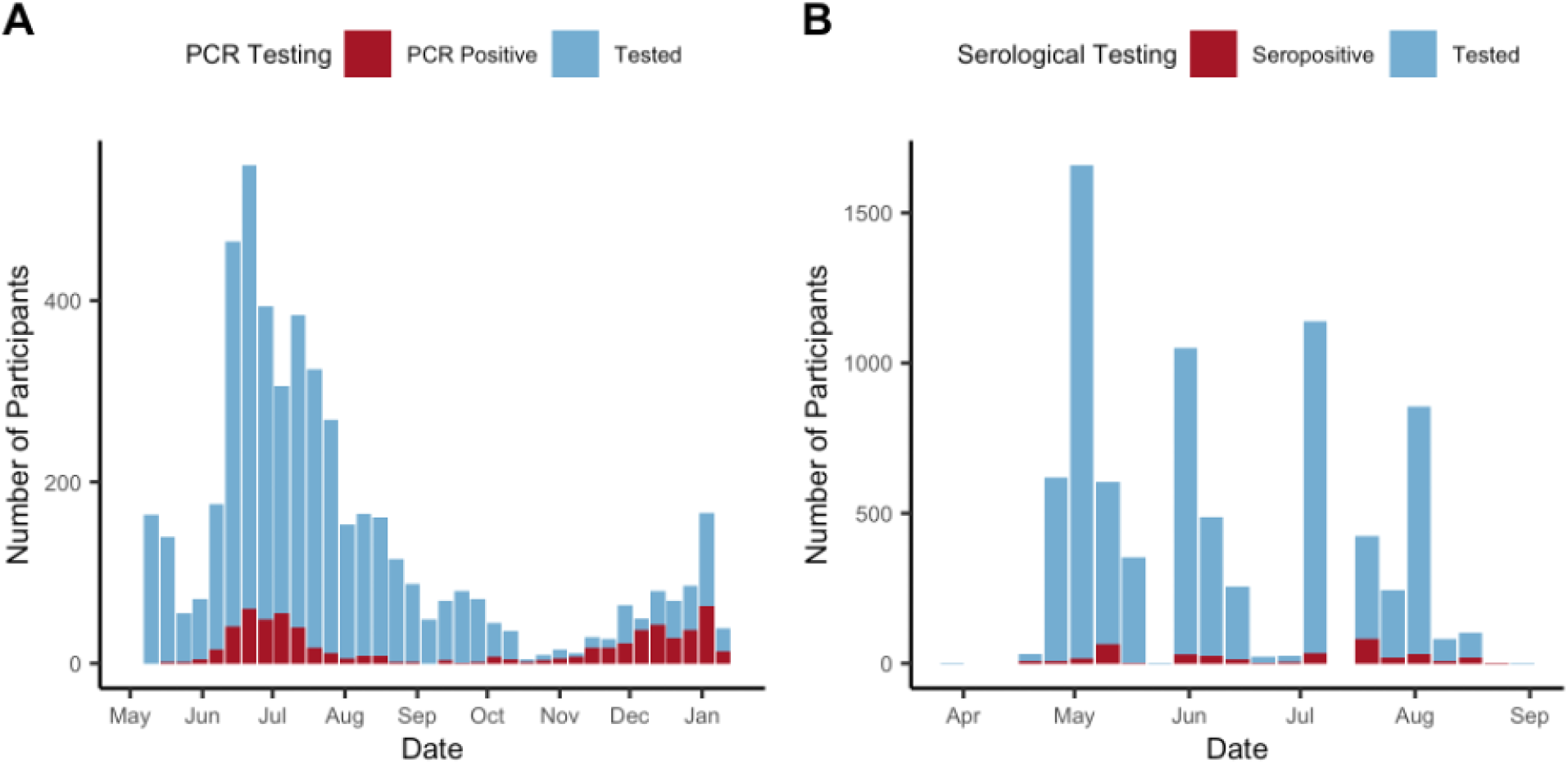
A) Number of PCR tests and PCR positive tests in the cohort between 5th April 2020 and 31st January 2021. B) Number of serological tests and seropositive tests between 29th March 2020 and 23rd August 2020.

### Risk of COVID-19 infection and reinfection

We estimated the odds ratio for SARS-CoV-2 reinfection using multivariable logistic regression. This required us to choose a cut-off week in order to define baseline seroprevalence and the subsequent observation period for PCR testing. To examine how our estimate for the odds ratio for reinfection varied depending on the cut-off week chosen, we repeated the analysis using every possible cut off week.

We estimated an adjusted odds ratio of 0·09 (95% CI: 0·005–0·48) for reinfection, with the week of 26th July 2020 as the optimal baseline time point (Figure 2), as this week fell after the first wave of infection and had the most precise odds ratio estimate. Odds ratio estimates using cut-off weeks in between the two waves of infection (between mid-July and mid-September 2020) ranged from 0.09 (95% CI: 0·005–0·48) to 0.25 (95% CI: 0.037–1.01).

**Figure 2:**
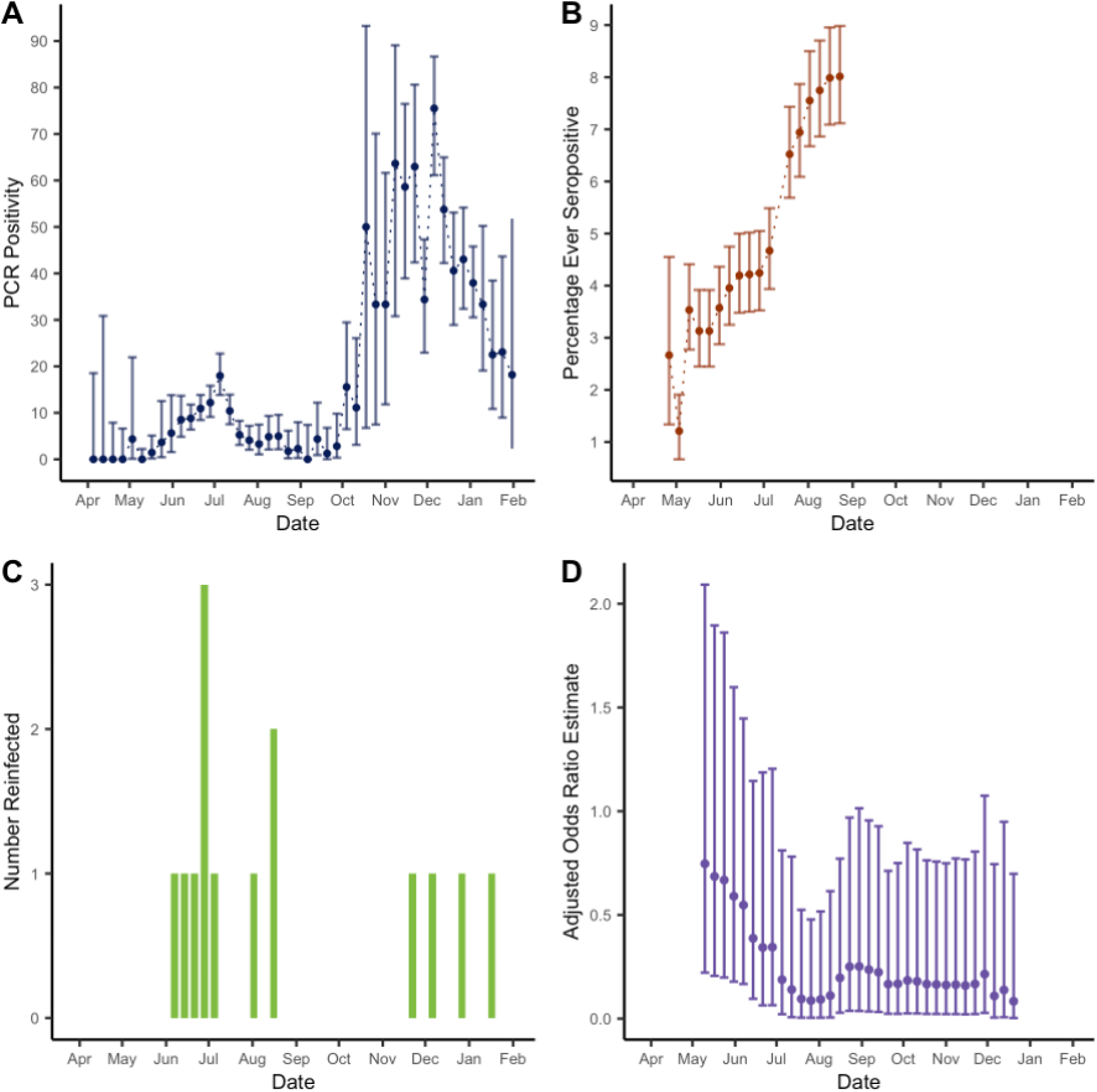
A) PCR positivity in the cohort between 5th April 2020 and 31st January 2021. B) Percentage ever seropositive in the cohort (number ever seropositive/ cumulative number enrolled) between 29th March 2020 and 23rd August 2020. Note that the percentage ever positive decreases initially as participants continue to be enrolled in the study. C) Number of possible reinfections in cohort over time (defined as a new positive PCR test more than 30 days after initial seropositive result). D) Odds ratio estimates comparing odds of reinfection in the seropositive group with odds of primary infection in the seronegative group, estimated using logistic regression and adjusted for race, ethnicity, state, job category and BMI. The estimates are presented with their associated 95% confidence intervals and with the cut-off week used to define baseline seroprevalence on the x-axis.

Unadjusted odds ratio estimates tended to overestimate the odds ratio for reinfection compared with primary infection, particularly when using early cut-off weeks (Figure 3). Notably, with early cut-off weeks the unadjusted analysis estimated a higher odds of reinfection compared to primary infection, albeit with wide confidence intervals. We hypothesise that individuals who are at higher risk of seroconversion (who would be included in analyses at earlier time thresholds) would also be at higher risk of later reinfection, giving a biased estimate of the effect of antibodies on subsequent infection for earlier cut-off weeks.

**Figure 3:**
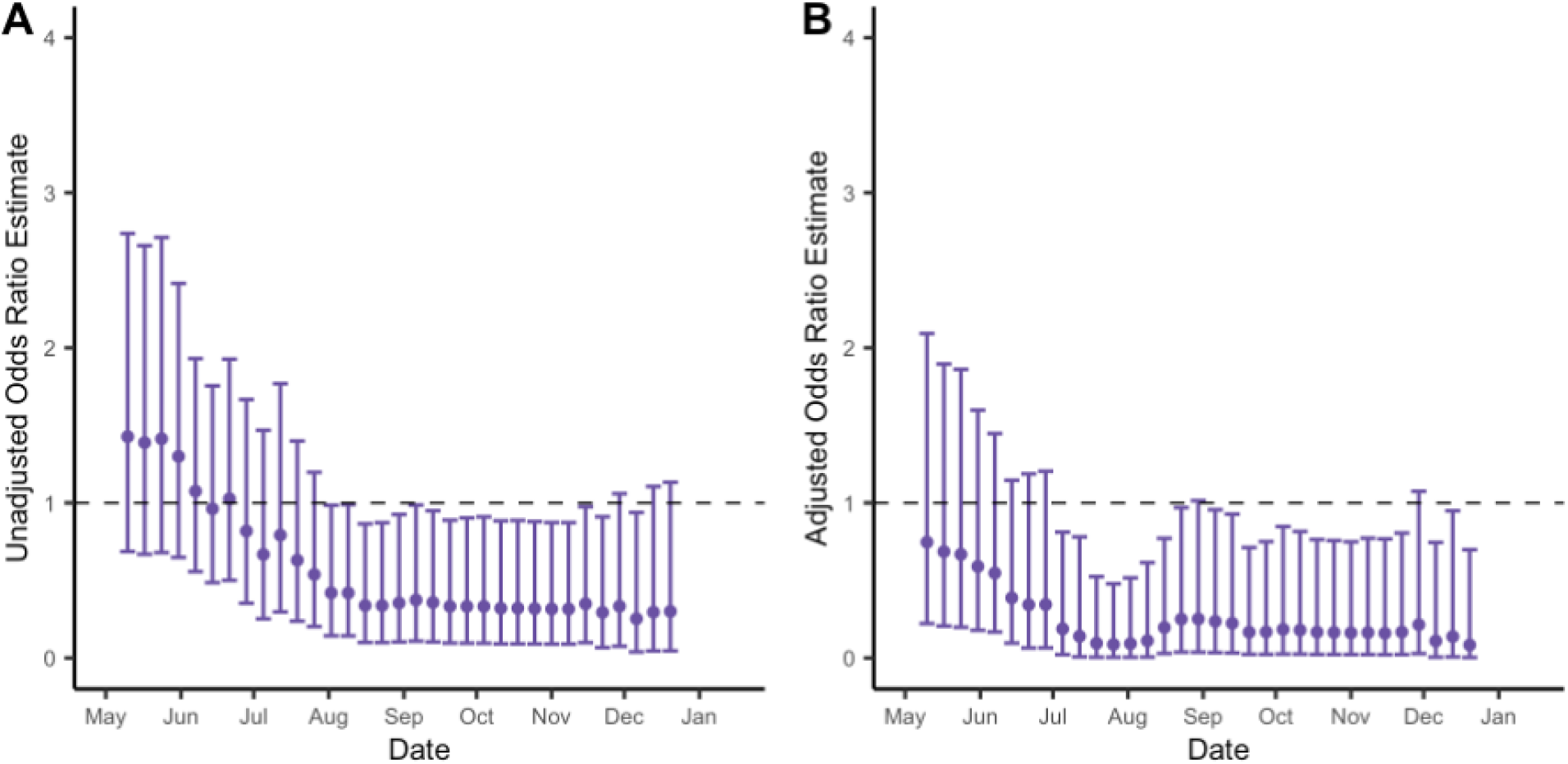
A) Unadjusted odds ratio estimates comparing odds of reinfection in the seropositive group with odds of primary infection in the seronegative group. The estimates are presented with their associated 95% confidence intervals and with the cut-off week used to define baseline seroprevalence on the x-axis. B) Odds ratio estimates comparing odds of reinfection in the seropositive group with odds of primary infection in the seronegative group, estimated using logistic regression and adjusted for race, ethnicity, state, job category and BMI. The estimates are presented with their associated 95% confidence intervals and with the cut-off week used to define baseline seroprevalence on the x-axis.

### Simulation using known underlying risk of infection

To further investigate how the precision of odds ratios estimates for reinfection varies depending on population-level epidemic dynamics, we conducted a simulation analysis using a known underlying infection risk distribution.

We used a probability distribution for risk of infection derived from PCR testing data from the study cohort. This was then scaled so that the overall cumulative risk reflected the level of seropositivity in the cohort by the end of the study period (8%). Considering a sample size of 2000 individuals over a period of 44 weeks, and a pre-set probability of reinfection given seropositivity of 0·15, we then re-estimated the corresponding risk ratios (Figure 4).

**Figure 4:**
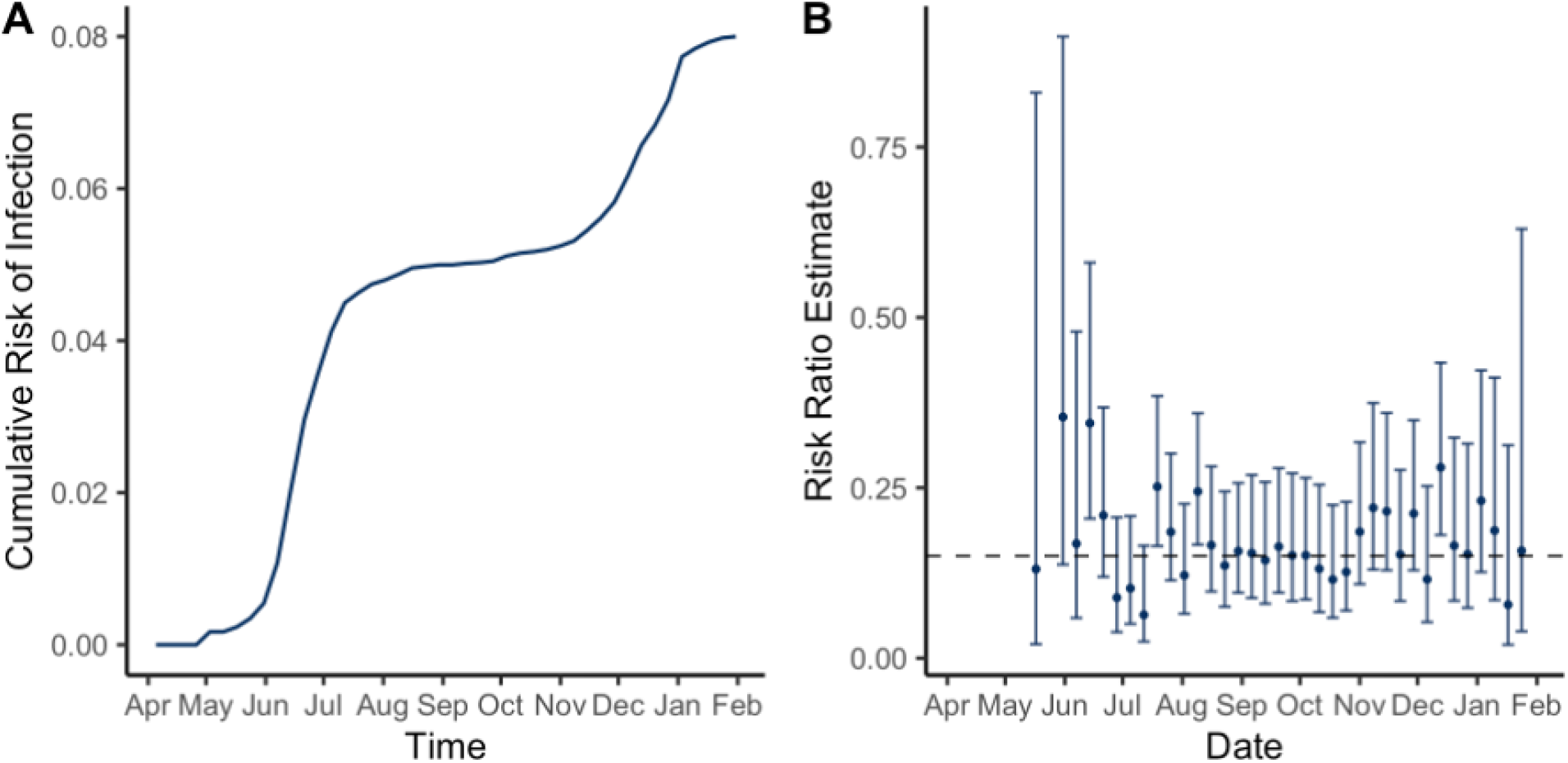
A) Cumulative risk of infection in the cohort used for simulation analysis. B) Risk ratio estimates comparing the risk of reinfection with the risk of primary infection. The estimates are presented with their associated 95% confidence intervals and with the cut-off week used to define baseline infection status on the x-axis. The dashed line represents the pre-set reinfection risk ratio of 0.15.

When considering this two-wave epidemic scenario, we found that the uncertainty in the estimated risk of reinfection was reduced in the middle of the simulation period (i.e. between the two ‘waves’ of infection risk). This supports our choice of optimal cut-off week for estimating the relative risk of reinfection in our study cohort, where we considered odds ratios given the need for an underlying regression analysis.

## Discussion

We identified 14 possible reinfections out of 309 seropositive individuals in this prospective seroepidemiological cohort. We estimated an adjusted odds ratio of 0·09 (95% CI: 0·005–0·48) for reinfection, with the week of 26th July 2020 as the optimal baseline time point in a study period between April 2020 and February 2021. This suggests that the presence of SARS-CoV-2 antibodies at baseline is associated with around 91% reduced odds of a subsequent PCR positive test. This provides further evidence that primary infection with SARS-CoV-2 results in protection against reinfection in the majority of individuals, at least over a sixth month time period. Our findings are consistent with estimates of 0·17 (95% CI 0·13-0·24) odds ratio [13] and 0·11 (0·03-0·44) incidence rate ratio [14] for healthcare workers, 0·18 (0·11-0·28) incidence rate ratio for military recruits [15] and 0·195 (95% CI 0·155–0·246) incidence rate ratio from a Danish population-level study [18].

Our analysis addressed two key sources of bias and uncertainty in estimating reinfection risk. First, confounders may inflate estimates; if a specific subset of the cohort is at higher risk of infection (e.g. due to underlying health conditions or increased risk of exposure), these participants will be more likely to be both initially seropositive and to have a subsequent reinfection. Second, the time period considered could increase uncertainty; defining the baseline seroprevalence at an early time point means few will be seropositive, whereas defining it at a later point means there is less time to observe possible reinfections. We accounted for these two factors by first using logistic regression to identify predictors of baseline seropositivity (i.e. infection risk), to calculate an adjusted odds ratio for reinfection. We then performed a sensitivity analysis to identify the optimum cut-off date to define baseline seroprevalence. We found that for a two-wave epidemic scenario, a cut-off week in the period in between the two waves of infection risk led to the most precise estimates of the odds ratio for reinfection. This demonstrates that the uncertainty surrounding estimates of reinfection risk will be sensitive to the study period chosen, relative to population-level epidemic dynamics.

There are several limitations to the underlying data that should be considered when interpreting these findings. This prospective cohort was recruited opportunistically from employees at one US company and is unlikely to be representative of the general population. However, as we did not identify any workplace outbreaks, transmission in this cohort is likely to be more reflective of community transmission than in health-care worker cohorts or other specialised populations. Additionally, we only considered possible reinfections (as opposed to probable or true reinfections). As possible reinfections did not meet a stringent case definition, such as confirmation through genomic sequencing, they may include cases of prolonged viral shedding following an initial infection. This would result in an overestimation of the odds ratio for reinfection and so our analysis reflects the minimum possible effect of antibodies on future SARS-CoV-2 infection risk.

As well as quantifying reinfection risk over a six-month period among a prospectively followed workplace population, our study highlights the importance of accounting for both individual-level heterogeneity in infection risk and population-level variation in epidemic dynamics when assessing the potential for reinfections.

## Methods

### Seroepidemiological cohort description

We used data from a seroepidemiological study of US employees at SpaceX, also described elsewhere [17]. In brief, this study involved 4411 employees from California, Florida, Texas and Washington State, with ages ranging from 18-71. All employees were invited to participate by email and there were no exclusion criteria. Study participants were offered SARS-CoV-2 IgG receptor-binding domain (RBD) antibody testing with an in-house ELISA assay with 82·4% sensitivity and 99·6% specificity [19]. Serological samples were taken during four rounds of testing between April - September 2020. A questionnaire including demographic, symptom and exposure information was conducted at enrolment, and with each round of serological testing.

Individuals continued to be enrolled throughout the study period, and around half of the total participants (48%) were tested at more than one time point. Participants occupied a range of job positions within SpaceX including office-based and factory-based jobs. Additionally, symptomatic and asymptomatic PCR testing were widely available for employees, with data available from April 2020 - January 2021. Both serology and PCR testing data were available for 1800 individuals.

### Statistical Analysis

To estimate the odds ratio for SARS-CoV-2 reinfection, we conducted multivariable logistic regression analysis investigating the association between baseline serological status and subsequent PCR test result, given a test was sought. Propensity to seek a PCR test did not appear to differ by baseline serostatus.

The choice of cut-off week used to define participants’ baseline seroprevalence and the subsequent observation period for PCR testing has important implications for the precision of the estimated odds ratio for reinfection. For instance, a cut-off week early in the study period will result in few seropositive individuals, while a cut-off week later in the study period leaves less time to observe subsequent PCR testing and detect possible reinfections. To identify the optimal cut-off week and assess how the choice of cut-off week affected estimates of the odds ratio for reinfection, we repeated the multivariable logistic regression for every possible cut-off week.

Potential confounding variables were selected a priori and included; age, sex, race, ethnicity, BMI, state, work location, job category, household size, history of chronic disease or history of smoking. We used logistic regression to identify predictors of baseline seropositivity (i.e. infection risk) using a forwards modelling strategy comparing the Akaike Information Criterion (AIC). Finally, we selected the optimally sized variable set to adjust for by comparing the AIC between models for the same cut-off week for different variable sets. Odds ratio estimates were adjusted for race, ethnicity, state, job category and BMI.

### Simulation analysis

We then conducted a simulation analysis to further investigate how the precision of odds ratio estimates for reinfection vary depending on the time point chosen to determine participants’ baseline seroprevalence and the subsequent window to monitor PCR infections and possible reinfections.

We simulated a two-wave epidemic scenario using a probability distribution for cumulative risk of infection derived from PCR testing data from the study cohort. This was scaled so the overall cumulative risk of infection reflected the overall level of seropositivity in the cohort (8%). We considered a sample size of 2000 individuals over a period of 44 weeks. For each time point the number of seropositive and seronegative individuals up to and including week was simulated, according to the background cumulative risk of infection. We then simulated how many of these seropositive and seronegative individuals would test PCR positive or negative after week, with a pre-set probability of reinfection of 0.15.

Finally, we re-estimated the risk ratio for reinfection for each cut-off time point and assessed the precision by comparing the estimated risk ratio over time with the ‘true’ ratio of 0.15.

Analysis was conducted in R version 4.0.3. Code to reproduce the figures and simulation analysis presented here can be found at https://github.com/EmilieFinch/covid-reinfection.

## Data Availability

Code to reproduce the figures and simulation analysis presented can be found at https://github.com/EmilieFinch/covid-reinfection.
The data that support the findings of this study are available on request by email. The data are not publicly available due to their containing information that could compromise the privacy of research participants.

## Funding

AJK was supported by Wellcome Trust (grant: 206250/Z/17/Z) and NIHR (NIHR200908). EF was funded by the Medical Research Council (grant number MR/N013638/1), RL was funded by a Royal Society Dorothy Hodgkin Fellowship. The work was also supported by the Translational Research Institute for Space Health through NASA Cooperative Agreement (NNX16AO69A), the Massachusetts Consortium on Pathogen Readiness (MassCPR), the NIH (3R37AI080289-11S1, R01AI146785, U19AI42790-01, U19AI135995-02, U19AI42790-01, 1U01CA260476-01), and the Musk Foundation.

We thank SpaceX employees Lindsay Chapman, Jordan Steinhart, Suzanne Siebert, and Kyle Meade for their valuable support, in addition to the dedication and commitment of the many SpaceX employees that volunteered to participate in this study.

## Competing interests

GA is a founder of Seromyx Systems Inc., a company developing platform technology that describes the antibody immune response. GA’s interests were reviewed and are managed by Massachusetts General Hospital in accordance with their conflict-of-interest policies. MJG, SB, DD, YH, JR, EP, BM, ASM, and ERM are employees of Space Exploration Technologies Corp. All other authors have declared that no conflict of interest exists.

## CMMID COVID-19 working group information

The following authors were part of the Centre for Mathematical Modelling of Infectious Disease COVID-19 Working Group. Each contributed in processing, cleaning and interpretation of data, interpreted findings, contributed to the manuscript, and approved the work for publication: Jiayao Lei, Sebastian Funk, Fiona Yueqian Sun, Amy Gimma, Emily S Nightingale, Graham Medley, Sam Abbott, Fabienne Krauer, Nicholas G. Davies, Mark Jit, Akira Endo, Oliver Brady, Anna M Foss, Yung-Wai Desmond Chan, Thibaut Jombart, Kevin van Zandvoort, Rosalind M Eggo, Yang Liu, Gwenan M Knight, Carl A B Pearson, Kaja Abbas, Katherine E. Atkins, Samuel Clifford, Mihaly Koltai, Yalda Jafari, Damien C Tully, Christopher I Jarvis, Kathleen O’Reilly, Nikos I Bosse, Kiesha Prem, Billy J Quilty, Simon R Procter, Rosanna C Barnard, William Waites, Ciara McCarthy, James D Munday, David Hodgson, W John Edmunds, Alicia Rosello, C Julian Villabona-Arenas, Hamish P Gibbs, Stefan Flasche, Timothy W Russell, Sophie R Meakin, Joel Hellewell, Naomi R Waterlow, Matthew Quaife, Frank G Sandmann.

Funding statements for the CMMID COVID-19 working group are as follows. KvZ: KvZ is supported by the UK Foreign, Commonwealth and Development Office (FCDO)/Wellcome Trust Epidemic Preparedness Coronavirus research programme (ref. 221303/Z/20/Z), and Elrha’s Research for Health in Humanitarian Crises (R2HC) Programme, which aims to improve health outcomes by strengthening the evidence base for public health interventions in humanitarian crises. The R2HC programme is funded by the UK Government (FCDO), the Wellcome Trust, and the UK National Institute for Health Research (NIHR). SC: Wellcome Trust (grant: 208812/Z/17/Z). FYS: NIHR EPIC grant (16/137/109). SFunk: Wellcome Trust (grant: 210758/Z/18/Z), NIHR (NIHR200908). GFM: NTD Modelling Consortium by the Bill and Melinda Gates Foundation (OPP1184344). YJ: LSHTM, DHSC/UKRI COVID-19 Rapid Response Initiative. SRM: Wellcome Trust (grant: 210758/Z/18/Z). WJE: European Commission (EpiPose 101003688), NIHR (NIHR200908). MQ: European Research Council Starting Grant (Action Number #757699); Bill and Melinda Gates Foundation (INV-001754). NRW: Medical Research Council (grant number MR/N013638/1). RME: HDR UK (grant: MR/S003975/1), MRC (grant: MC_PC 19065), NIHR (grant: NIHR200908). NGD: UKRI Research England; NIHR Health Protection Research Unit in Immunisation (NIHR200929); UK MRC (MC_PC_19065). JYL: Bill & Melinda Gates Foundation (INV-003174). MK: Foreign, Commonwealth and Development Office / Wellcome Trust. FK: Innovation Fund of the Joint Federal Committee (Grant number 01VSF18015), Wellcome Trust (UNS110424). DCT: No funding declared. JDM: Wellcome Trust (grant: 210758/Z/18/Z). AS: No funding declared. AMF: No funding declared. KP: Gates (INV-003174), European Commission (101003688). SFlasche: Wellcome Trust (grant: 208812/Z/17/Z). SA: Wellcome Trust (grant: 210758/Z/18/Z). BJQ: This research was partly funded by the National Institute for Health Research (NIHR) (16/137/109 & 16/136/46) using UK aid from the UK Government to support global health research. The views expressed in this publication are those of the author(s) and not necessarily those of the NIHR or the UK Department of Health and Social Care. BJQ is supported in part by a grant from the Bill and Melinda Gates Foundation (OPP1139859). TJ: RCUK/ESRC (grant: ES/P010873/1); UK PH RST; NIHR HPRU Modelling & Health Economics (NIHR200908). AR: NIHR (grant: PR-OD-1017-20002). GMK: UK Medical Research Council (grant: MR/P014658/1). MJ: Gates (INV-003174, INV-016832), NIHR (16/137/109, NIHR200929, NIHR200908), European Commission (EpiPose 101003688). YL: Gates (INV-003174), NIHR (16/137/109), European Commission (101003688). JW: NIHR Health Protection Research Unit and NIHR HTA. JH: Wellcome Trust (grant: 210758/Z/18/Z). KO’R: Bill and Melinda Gates Foundation (OPP1191821). YWDC: No funding declared. TWR: Wellcome Trust (grant: 206250/Z/17/Z). CIJ: Global Challenges Research Fund (GCRF) project ‘RECAP’ managed through RCUK and ESRC (ES/P010873/1). SRP: Bill and Melinda Gates Foundation (INV-016832). AE: The Nakajima Foundation. ESN: Gates (OPP1183986). NIB: Health Protection Research Unit (grant code NIHR200908). CJVA: European Research Council Starting Grant (Action number 757688). FGS: NIHR Health Protection Research Unit in Modelling & Health Economics, and in Immunisation. AG: European Commission (EpiPose 101003688). KA: Bill & Melinda Gates Foundation (OPP1157270, INV-016832). WW: MRC (grant MR/V027956/1). KEA: European Research Council Starting Grant (Action number 757688). RCB: European Commission (EpiPose 101003688). PK: This research was partly funded by the Royal Society under award RP\EA\180004, European Commission (101003688), Bill & Melinda Gates Foundation (INV-003174). HPG: This research was produced by CSIGN which is part of the EDCTP2 programme supported by the European Union (grant number RIA2020EF-2983-CSIGN). The views and opinions of authors expressed herein do not necessarily state or reflect those of EDCTP. This research is funded by the Department of Health and Social Care using UK Aid funding and is managed by the NIHR. The views expressed in this publication are those of the author(s) and not necessarily those of the Department of Health and Social Care (PR-OD-1017-20001). CABP: CABP is supported by the Bill & Melinda Gates Foundation (OPP1184344) and the UK Foreign, Commonwealth and Development Office (FCDO)/Wellcome Trust Epidemic Preparedness Coronavirus research programme (ref. 221303/Z/20/Z). OJB: Wellcome Trust (grant: 206471/Z/17/Z). WJE: European Commission (EpiPose 101003688), NIHR (NIHR200908, MRC: MC_PC_19065). CIJ: Global Challenges Research Fund (GCRF) project ‘RECAP’ managed through RCUK and ESRC (ES/P010873/1). NGD: UKRI Research England; NIHR Health Protection Research Unit in Immunisation (NIHR200929); UK MRC (MC_PC_19065).

## Notes

### Author Declarations

The study protocol was approved by the Western Institutional Review Board. The use of de-identified data and biological samples was approved by the Mass General Brigham Healthcare Institutional Review Board. All participants provided written informed consent.

## References

1. Baker RE, Yang W, Vecchi GA, Metcalf CJE, Grenfell BT. Susceptible supply limits the role of climate in the early SARS-CoV-2 pandemic. Science. 2020 Jul 17;369(6501):315–9.

2. Sabino EC, Buss LF, Carvalho MPS, Prete CA, Crispim MAE, Fraiji NA, et al. Resurgence of COVID-19 in Manaus, Brazil, despite high seroprevalence. The Lancet. 2021 Feb 6;397(10273):452–5.

3. Seow J, Graham C, Merrick B, Acors S, Pickering S, Steel KJA, et al. Longitudinal observation and decline of neutralizing antibody responses in the three months following SARS-CoV-2 infection in humans. Nat Microbiol. 2020 Dec;5(12):1598–607.

4. Long Q-X, Liu B-Z, Deng H-J, Wu G-C, Deng K, Chen Y-K, et al. Antibody responses to SARS-CoV-2 in patients with COVID-19. Nat Med. 2020 Jun;26(6):845–8.

5. Gudbjartsson DF, Norddahl GL, Melsted P, Gunnarsdottir K, Holm H, Eythorsson E, et al. Humoral Immune Response to SARS-CoV-2 in Iceland. N Engl J Med. 2020 Oct 29;383(18):1724–34.

6. Ibarrondo FJ, Fulcher JA, Goodman-Meza D, Elliott J, Hofmann C, Hausner MA, et al. Rapid Decay of Anti–SARS-CoV-2 Antibodies in Persons with Mild Covid-19. N Engl J Med [Internet]. 2020 Jul 21 [cited 2021 Apr 28]; Available from: https://www.nejm.org/doi/10.1056/NEJMc2025179

7. Chia WN, Zhu F, Ong SWX, Young BE, Fong S-W, Bert NL, et al. Dynamics of SARS-CoV-2 neutralising antibody responses and duration of immunity: a longitudinal study. Lancet Microbe [Internet]. 2021 Mar 23 [cited 2021 Apr 28];0(0). Available from: https://www.thelancet.com/journals/lanmic/article/PIIS2666-5247(21)00025-2/abstract

8. Duysburgh E, Mortgat L, Barbezange C, Dierick K, Fischer N, Heyndrickx L, et al. Persistence of IgG response to SARS-CoV-2. Lancet Infect Dis. 2021 Feb 1;21(2):163–4.

9. Dan JM, Mateus J, Kato Y, Hastie KM, Yu ED, Faliti CE, et al. Immunological memory to SARS-CoV-2 assessed for up to 8 months after infection. Science [Internet]. 2021 Feb 5 [cited 2021 Apr 28];371(6529). Available from: https://science.sciencemag.org/content/371/6529/eabf4063

10. Ward H, Cooke G, Atchison C, Whitaker M, Elliott J, Moshe M, et al. Declining prevalence of antibody positivity to SARS-CoV-2: a community study of 365,000 adults. medRxiv. 2020 Oct 27;2020.10.26.20219725.

11. Long Q-X, Tang X-J, Shi Q-L, Li Q, Deng H-J, Yuan J, et al. Clinical and immunological assessment of asymptomatic SARS-CoV-2 infections. Nat Med. 2020 Aug;26(8):1200–4.

12. To KK-W, Hung IF-N, Ip JD, Chu AW-H, Chan W-M, Tam AR, et al. Coronavirus Disease 2019 (COVID-19) Re-infection by a Phylogenetically Distinct Severe Acute Respiratory Syndrome Coronavirus 2 Strain Confirmed by Whole Genome Sequencing. Clin Infect Dis [Internet]. 2020 Aug 25 [cited 2021 Apr 28];(ciaa1275). Available from: https://doi.org/10.1093/cid/ciaa1275

13. Hall VJ, Foulkes S, Charlett A, Atti A, Monk EJ, Simmons R, et al. SARS-CoV-2 infection rates of antibody-positive compared with antibody-negative health-care workers in England: a large, multicentre, prospective cohort study (SIREN). The Lancet [Internet]. 2021 Apr 9 [cited 2021 Apr 14];0(0). Available from: https://www.thelancet.com/journals/lancet/article/PIIS0140-6736(21)00675-9/abstract

14. Lumley SF, O’Donnell D, Stoesser NE, Matthews PC, Howarth A, Hatch SB, et al. Antibody Status and Incidence of SARS-CoV-2 Infection in Health Care Workers. N Engl J Med. 2021 Feb 11;384(6):533–40.

15. Letizia AG, Ge Y, Vangeti S, Goforth C, Weir DL, Kuzmina NA, et al. SARS-CoV-2 seropositivity and subsequent infection risk in healthy young adults: a prospective cohort study. medRxiv. 2021 Jan 29;2021.01.26.21250535.

16. Addetia A, Crawford KHD, Dingens A, Zhu H, Roychoudhury P, Huang M-L, et al. Neutralizing Antibodies Correlate with Protection from SARS-CoV-2 in Humans during a Fishery Vessel Outbreak with a High Attack Rate. J Clin Microbiol [Internet]. 2020 Oct 21 [cited 2021 Mar 3];58(11). Available from: https://jcm.asm.org/content/58/11/e02107-20

17. Bartsch YC, Fischinger S, Siddiqui SM, Chen Z, Yu J, Gebre M, et al. Discrete SARS-CoV-2 antibody titers track with functional humoral stability. Nat Commun. 2021 Dec;12(1):1018.

18. Hansen CH, Michlmayr D, Gubbels SM, Mølbak K, Ethelberg S. Assessment of protection against reinfection with SARS-CoV-2 among 4 million PCR-tested individuals in Denmark in 2020: a population-level observational study. The Lancet [Internet]. 2021 Mar 17 [cited 2021 Mar 24];0(0). Available from: https://www.thelancet.com/journals/lancet/article/PIIS0140-6736(21)00575-4/abstract

19. Roy V, Fischinger S, Atyeo C, Slein M, Loos C, Balazs A, et al. SARS-CoV-2-specific ELISA development. J Immunol Methods. 2020 Sep 1;484–485:112832.

